# Pre-pandemic SARS-CoV-2 serological reactivity in rural malaria-experienced Cambodians

**DOI:** 10.1101/2021.09.27.21264000

**Authors:** Jessica Manning, Irfan Zaidi, Chanthap Lon, Luz Angela Rosas, Jae-Keun Park, Aiyana Ponce, Jennifer Bohl, Sophana Chea, Maria Karkanitsa, Sokunthea Sreng, Huy Rekol, Char Meng Chour, Dominic Esposito, Jeffery K. Taubenberger, Matthew J. Memoli, Kaitlyn Sadtler, Patrick E. Duffy, Fabiano Oliveira

## Abstract

Greater Mekong inhabitants are exposed to pathogens, zoonotic and otherwise, that may influence SARS-CoV-2 seroreactivity. A pre-pandemic (2005 to 2011) serosurvey of from 528 malaria-experienced Cambodians demonstrated higher-than-expected (up to 13.8 %) positivity of non-neutralizing IgG to SARS-CoV-2 spike and RBD antigens. These findings have implications for interpreting large-scale serosurveys.

**Article Summary Line:** In the pre-COVID19 pandemic years of 2005 to 2011, malaria experienced Cambodians from rural settings had higher-than-expected seroreactivity to SARS-CoV-2 spike and receptor binding domain proteins.

## Text

There are no published SARS-CoV-2 serosurveys in the Greater Mekong Subregion (GMS), aside from screening healthcare workers in two urban hospital-based settings (1,2). These antibody-based studies are necessary to estimate at-risk populations and to direct disease containment measures; however, prior to informing public health decisions, serological assays require careful country-specific calibration as several regions report fluctuating results or high background reactivity in different populations (3–5). This variability may be attributable to the myriad serological assays, the hypothesized cross-reactivity from ‘common cold’-type respiratory coronaviruses (6), prior *Plasmodium* infections (7–9), or previously uncharacterized betacoronaviruses in wildlife populations found in the rural GMS (10–12). While countless serological SARS-CoV-2 investigations are currently underway, it is prudent to consider how the broad pathogen diversity in the GMS may influence estimations of SARS-CoV-2 seroprevalence.

## The Study

We tested sera or plasma from 528 malaria-infected Cambodian individuals sampled from 2005 to 2011 (prior to the presumed emergence of SARS-CoV-2 in 2019) for IgG antibodies reactive to SARS-CoV-2 spike and receptor binding domain (RBD) proteins via an optimized enzyme-linked immunosorbent assay (ELISA)(13,14).

Because six other coronaviruses (OC43, HKU1, 229E, NL63, SARS CoV-1, and MERS viruses) possess structural proteins capable of infecting humans, we selected a highly specific ELISAs for the aforementioned SARS-CoV-2 structural proteins (13,14). Compared to other coronaviruses, SARS-CoV-2 shows varying levels of spike protein sequence homology, with the highest for SARS-CoV-1 (76% identity; 87% similarity) and the lowest for the ‘common cold’ coronavirus HKU1 (29% identity; 40% similarity)(13). Reactivity to both spike and RBD antigens above cutoff values is required for a positive test with reported sensitivity and specificity of 100% (95% CI: 92.9 to 100) and 100% (95% CI: 98.8 to 100)(13,14). Pre-pandemic samples had levels above the set cutoffs for SARS-CoV-2 spike and RBD antigens (Figure 1) varying from 4.4 to 13.8% positivity to both SARS-CoV-2 spike and RBD depending on which cutoff values (calibrated for the Mali or US populations) are used for this specific assay (4,13,14) (Table 1, Figure 1, Appendix Table 1).

**Table 1.**
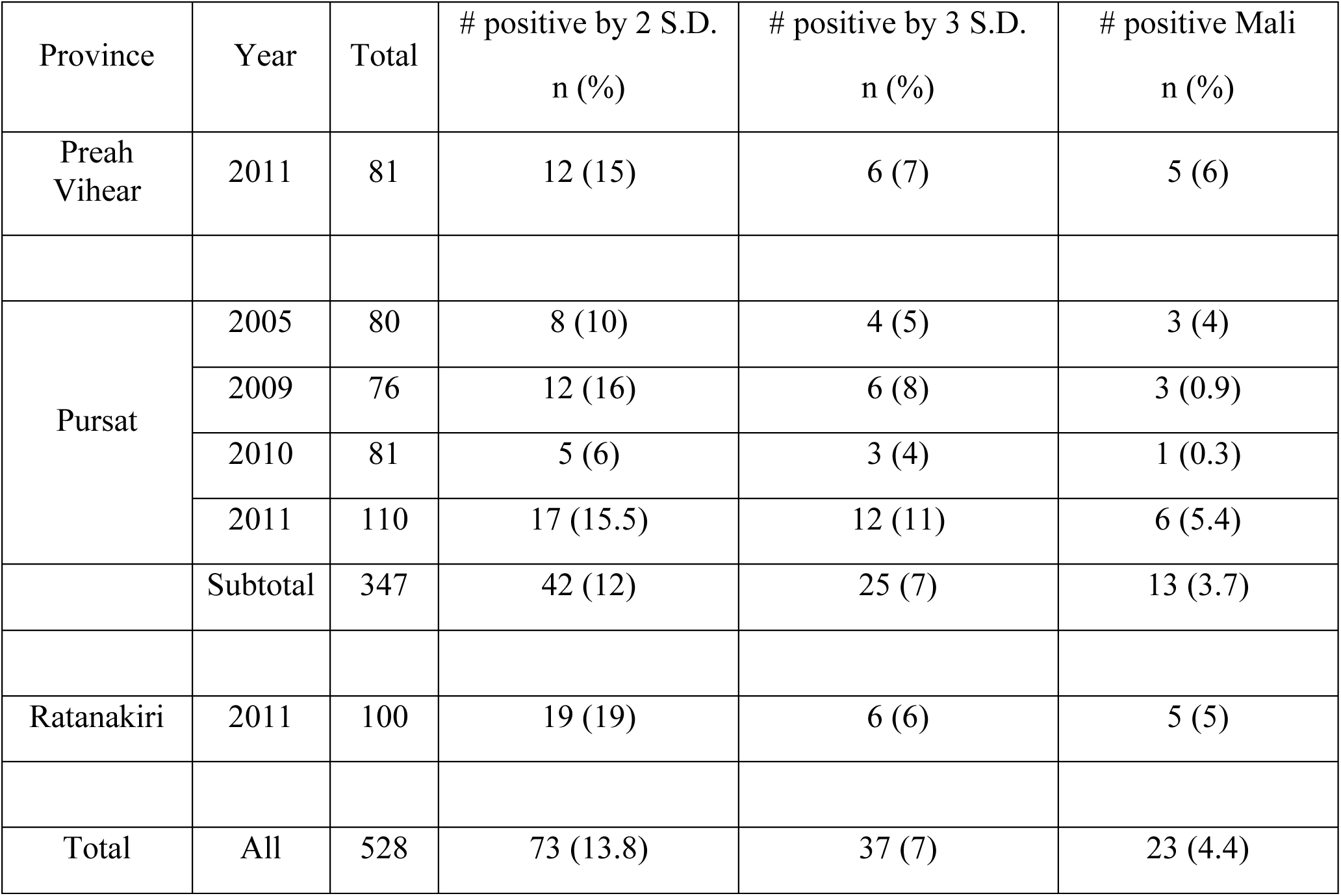
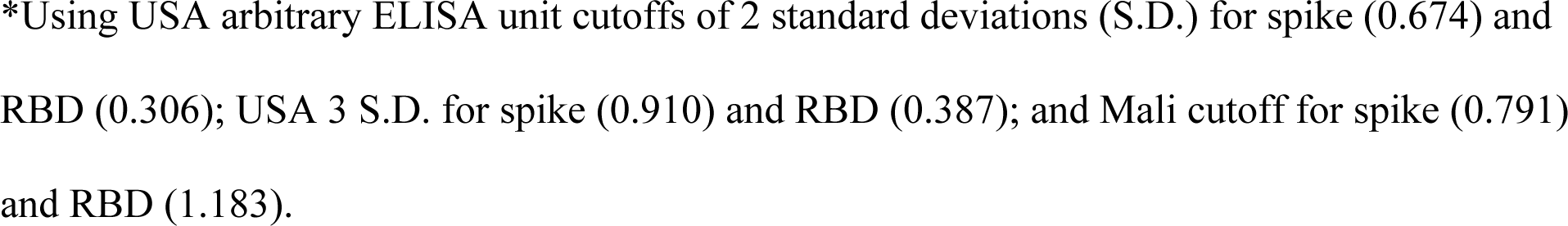
SARS-CoV-2 ELISA results by cutoff values in three Cambodian provinces from 2005 to 2011.

| Province | Year | Total | # positive by 2 S.D.<br>n (%) | # positive by 3 S.D.<br>n (%) | # positive Mali<br>n (%) |
| --- | --- | --- | --- | --- | --- |
| Preah Vihear | 2011 | 81 | 12 (15) | 6 (7) | 5 (6) |
| Pursat | 2005 | 80 | 8 (10) | 4 (5) | 3 (4) |
|  | 2009 | 76 | 12 (16) | 6 (8) | 3 (0.9) |
|  | 2010 | 81 | 5 (6) | 3 (4) | 1 (0.3) |
|  | 2011 | 110 | 17 (15.5) | 12 (11) | 6 (5.4) |
|  | Subtotal | 347 | 42 (12) | 25 (7) | 13 (3.7) |
| Ratanakiri | 2011 | 100 | 19 (19) | 6 (6) | 5 (5) |
| Total | All | 528 | 73 (13.8) | 37 (7) | 23 (4.4) |

\*Using USA arbitrary ELISA unit cutoffs of 2 standard deviations (S.D.) for spike (0.674) and RBD (0.306); USA 3 S.D. for spike (0.910) and RBD (0.387); and Mali cutoff for spike (0.791) and RBD (1.183).

**Figure 1.**
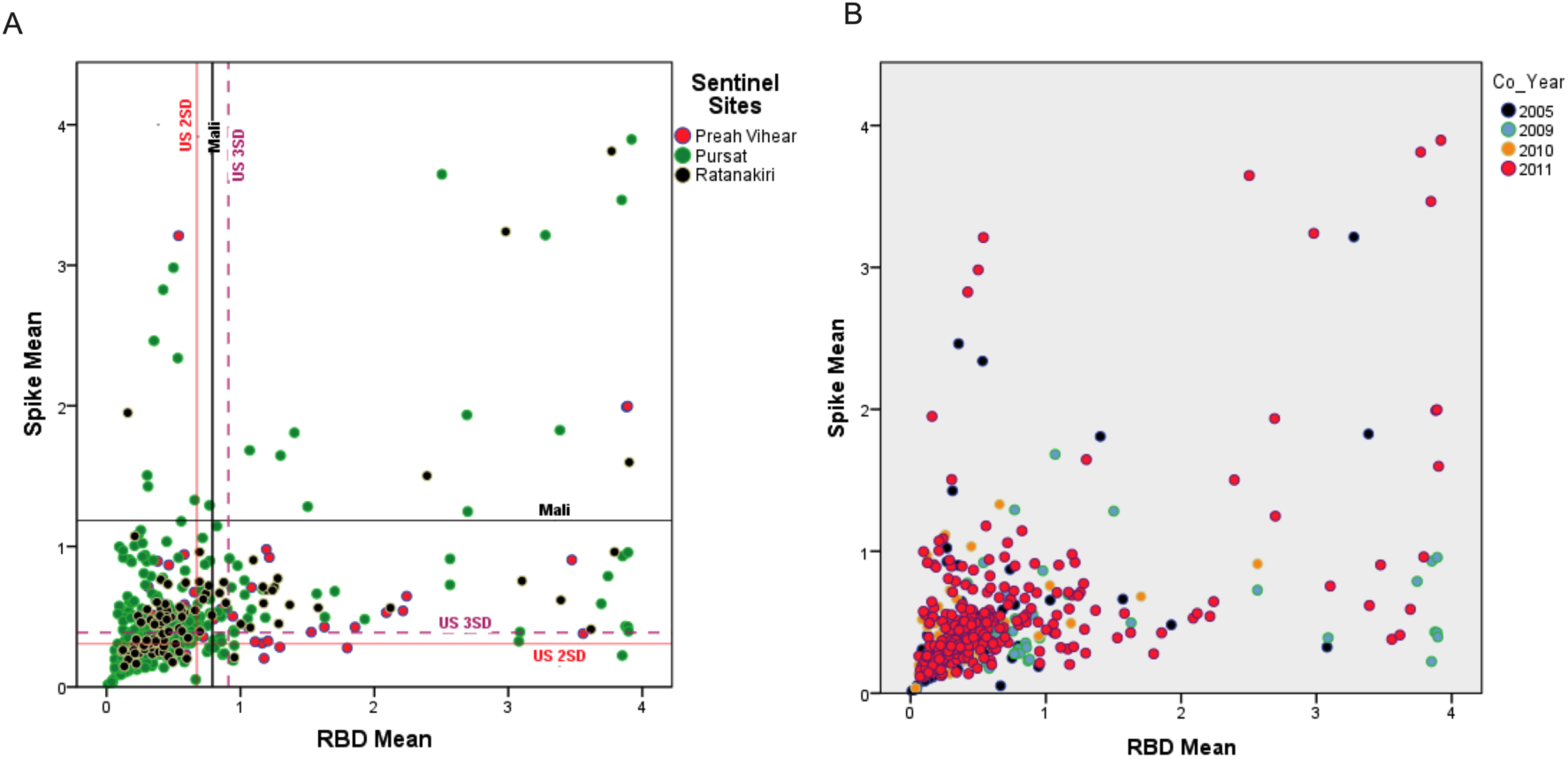
Mean antibody intensity in arbitrary ELISA units to Spike and Receptor Binding Domain (RBD) in pre-pandemic, malaria-positive Cambodian sera samples colored by province in (A) as Preah Vihear (pink), Pursat (green), Ratanakiri (black); and by year in (B) as 2005 (purple), 2009 (turquoise), 2010 (orange), and 2011 (pink).

To test whether the higher-than-expected positivity was an artifact of our in-house ELISA assay, we assayed a subset of samples with a commercially validated SARS-CoV-2 Spike S1-RBD IgG ELISA Detection commercial Kit (Genscript). Of the 24 individuals who were seronegative and 11 seropositives in the in-house assay, 18 tested negative and 9 tested positive in the commercial test, respectively, yielding an overall concordance of 77.1% between assays (Appendix Table 2). This inconsistency may be explained by the stringency of the in-house assay that tests both spike and RBD versus the commercial kit that tests for RBD only; nevertheless, the higher-than-expected positivity is observed in both assays. Since common cold coronaviruses do circulate in Cambodia, but no documented SARS-CoV-1 or MERS, we tested a subset of the cohort for IgG antibodies to HKU1 and OC43. Reactivity between subjects was comparable despite SARS-CoV-2 serostatus (Figure 2A).

**Figure 2.**
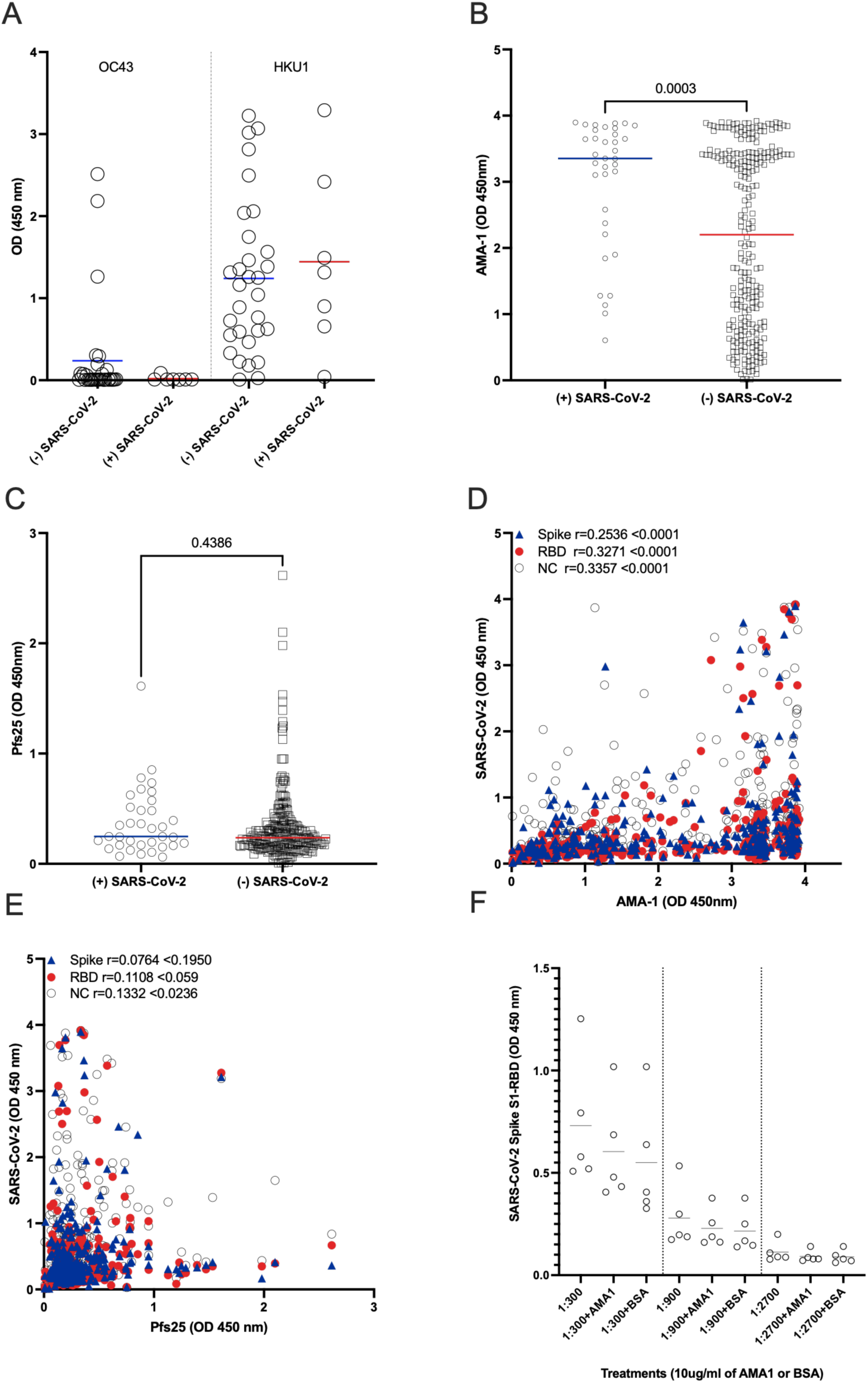
Mean antibody levels to (A) Common cold OC43 and HKU1 viruses, (B) *Plasmodium falciparum* Apical Membrane Antigen1 (AMA-1) and (C) Plasmodium falciparum Pfs25 protein (Pfs25) by SARS-CoV-2 serosurvey statuses. (D-E) Correlation of mean IgG antibody levels of (C) AMA-1 or (D) Pfs25 against Spike (blue triangles), Receptor Binding Domain (RBD – red circles) and Nucleocapsid (NC – open circles) IgG antibody levels in pre-pandemic, malaria-positive Cambodian sera samples (F) OD levels of RBD protein after preincubation of sera with 10mg/ml of AMA1 or BSA.

We further tested 289 subjects to assess if there was a relationship between antibodies to *Plasmodium spp*. and SARS-CoV-2 proteins using two known malarial antigens, *Plasmodium falciparum* Apical Membrane Antigen1 (AMA-1; highly immunogenic and an indicator of parasite exposure) and *Plasmodium falciparum* Pfs25 protein (Pfs25; poorly immunogenic and expressed only during the mosquito stages of parasite development (4)) (Figure 2B–E). Notably, when we split subjects by their SARS-CoV-2 serostatus, we detected significantly higher levels of AMA-1 antibodies in SARS-CoV-2 seropositive individuals (mean AMA-1 antibody level 3.0 versus 2.1 respectively; p=0.0003)(Figure 2B). As expected, there was no difference in antibody levels to Pfs25 with regard to SARS-CoV-2 seropositivity (Figure 2C). A weak but statistically significant positive correlation was detected between spike and RBD with AMA-1 IgG antibodies (Figure 2D). This finding corroborates recent observations that higher SARS-CoV-2 seroreactivity by ELISA or rapid tests is detected in individuals from malaria-endemic areas, expanding previous observations to include Southeast Asia (7–9). We also evaluated the samples for seroreactivity against the nucleocapsid (NC) protein that also positively correlated with the AMA-1 IgG antibodies. Only NC antibodies were weakly correlated to Pfs25 antibodies, which reinforces the argument for non-specific reactivity of NC (Figure 2E). Pre-incubation with 10 mg/ml of AMA-1 or BSA had no significant effect in the reactivity to SARS-CoV-2 Spike S1-RBD (Figure 2F). Taken together with studies elsewhere, *Plasmodium spp*. exposure may contribute to SARS-CoV-2 malaria-related background reactivity. This reactivity could be attributed to immune responses to other *Plasmodium spp*. proteins, polyclonal B-cell activation during infection, or interaction with the sialic acid moiety on N-linked glycans of the SARS-CoV-2 spike protein (7,8). Of note, SARS-CoV-2 Spike proteins used in the aforementioned assays were produced in HEK293 mammalian cells and likely have comparable glycosylation patterns. Elsewhere, malaria-induced cross reactivity, in pre-pandemic malaria-experienced African samples, was mitigated by modification of two commercial assays by the addition of a urea wash (7).

To understand the functionality of the antibodies present, we took a subset (n=21) of the samples with the highest reactivity to SARS-CoV-2 total IgG and performed neutralization assays as described in Appendix Figure 1. No neutralizing activity was identified despite high levels of antibodies reacting to both spike and RBD proteins. Identical results were obtained using surrogate virus neutralization test (sVNT) targeting the RBD’s interaction with the host cell receptor ACE2 (Genscript) (Appendix Table 3)(15). Both SARS-CoV-2 infection and vaccination can trigger high levels of non-neutralizing antibodies while neutralizing antibodies aimed primarily at the RBD seem to wane faster and remain at low titers (15). Plausibly, the cross-reactive non-functional antibodies to SARS-CoV-2 were raised during an infection by *Plasmodium spp*. (7), but we cannot discard the hypothesis that the presence of non-neutralizing SARS-CoV-2-reactive antibodies in pre-pandemic sera may be linked to betacoronaviruses’ ability in general to evade immune recognition via their complex surfaces (15,16). A limitation in understanding the assays’ specificity in pre-pandemic and from convalescent samples from confirmed SARS-CoV-2 infection in present-day Cambodian patients.

## Conclusions

We found in a widely-used, highly specific, and validated enzyme linked-immunosorbent assay that approximately 4 – 14% of pre-pandemic Cambodian sera samples were positive for non-neutralizing antibodies to SARS-CoV-2 spike and RBD antigens using various standardized optical density cut-off values (4,13,14). A relationship was noted between increased SARS-CoV-2 seroreactivity and anti-malarial humoral immunity as was also recently shown in Africa (7). The plausibility of regular spillover events, or simply increased exposure to uncharacterized betacoronaviruses, as a reason for SARS-CoV-2 cross-reactivity is also increased in these high-risk settings for zoonotic disease transmission, given agricultural and dietary practices such as bat guano collection and consumption of wild meats (10–12). Given 50 to 80% of GMS residents are classified as rural, careful calibration of serological assays targeting SARS-CoV-2 will be necessary in national and subnational serosurveys. While neutralization assays with live virus are often considered the gold standard for their specificity, they are cost-prohibitive for large-scale serosurveys. The use of competition ELISA assays such sVNT targeting the RBD-ACE2 blockade may be an attractive option for populations at high-risk for zoonotic exposures in resource-scarce settings without BSL-3 facilities.

## Ethics Statement

These malaria research studies (NCT00341003, NCT00663546, NCT01350856) that collected de-identified, anonymized sera or plasma samples for future use, as done in this study published here, was approved by the Institutional Review Boards in the USA (National Institute of Allergy and Infectious Diseases Institutional Review Board) and in Cambodia (National Ethics Committee on Human Research; FWA #10451, IRB #3143).

## Data Availability

Data is available in appendix and also upon request.

## Acknowledgments

We thank the participants of the studies in Pursat, Ratanakiri, and Preah Vihear provinces and the original study protocol staff. This research was supported by the Intramural Research Program of the NIH, National Institute of Allergy and Infectious Diseases.

Data is available in appendix and also upon request.

