## Appendix for "Pre-pandemic SARS-CoV-2 serological reactivity in rural malaria-experienced Cambodians"

Appendix Table 1. Participants' SARS-CoV-2 Seroreactivity by Site, Year, Gender and Age.

| Site | Samples | Year | Males | Female | <10 years | 10-17 years | >18 years |
|---|---|---|---|---|---|---|---|
| Preah Vihear | 81 (15) | 2011 | 51 (9.7) | 30 (5.7) | 7 (1.6) | 27 (6.0) | 47 (10.5) |
| Pursat | 347 (66) | 2005 | 261 (49.4) | 86 (16.3) | 20 (4.5) | 71 (15.9) | 256 (57.3) |
| | | 2009 | | | | | |
| | | 2010 | | | | | |
| | | 2011 | | | | | |
| Ratanakiri* | 100 (19) | 2011 | 11 (2.1) | 8 (1.5) | 6 (1.3) | 7 (1.6) | 6 (1.3) |
| **Total** | **528** | **528** | **323 (61.2)** | **124 (23.5)** | **33 (7.4)** | **105 (23.5)** | **309 (69.1)** |

All values n (%) unless otherwise noted.

*81 samples missing gender and age data

Appendix Table 2. Seropositivity status by two SARS-CoV-2 IgG assays.

| Subject number | Test 1 Spike / RBD 2 S.D. cutoff | Mean RBD | Mean Spike | Spike S1-RBD commercial<br><br>Mean OD450 / Cutoff > 1 | Mean OD450 | Mean OD450 / Cutoff > 1 |
|---|---|---|---|---|---|---|
| 1 | Negative | 0.2157 | 0.2637 | Negative | 0.0000 | 0.0000 |
| 2 | Negative | 0.0813 | 0.1241 | Negative | 0.0005 | 0.0041 |
| 3 | Negative | 0.0847 | 0.3078 | Negative | 0.0220 | 0.1812 |
| 4 | Negative | 0.0480 | 0.0532 | Negative | 0.0077 | 0.0630 |
| 5 | Negative | 0.0854 | 0.5203 | Negative | 0.0081 | 0.0663 |
| 6 | Negative | 0.0616 | 0.1983 | Negative | 0.0221 | 0.1820 |
| 7 | **Negative** | **0.0613** | **0.1358** | **Positive** | **0.2514** | **2.0708** |
| 8 | Negative | 0.0369 | 0.0361 | Negative | 0.0215 | 0.1767 |
| 9 | Positive | 3.7444 | 0.7881 | Positive | 1.0889 | 8.9691 |
| 10 | Negative | 0.1912 | 0.2379 | Negative | 0.0112 | 0.0923 |
| 11 | **Negative** | **3.8750** | **0.4312** | **Positive** | **0.6211** | **5.1161** |
| 12 | **Negative** | **3.8917** | **0.4280** | **Positive** | **0.6598** | **5.4349** |
| 13 | **Negative** | **3.8971** | **0.3964** | **Positive** | **0.7262** | **5.9815** |
| 14 | **Negative** | **3.0869** | **0.3910** | **Positive** | **0.4061** | **3.3451** |
| 15 | **Positive** | **3.8931** | **0.9565** | **Negative** | **0.0825** | **0.6796** |
| 16 | **Positive** | **3.8532** | **0.9306** | **Negative** | **0.0925** | **0.7615** |
| 17 | Positive | 2.5646 | 0.7263 | Positive | 0.2392 | 1.9703 |
| 18 | Positive | 3.8824 | 1.9920 | Positive | 0.4771 | 3.9296 |

| | | | | | | |
|---|---|---|---|---|---|---|
| 19 | Positive | 3.4743 | 0.9039 | Positive | 0.5926 | 4.8814 |
| 20 | **Negative** | **2.2409** | **0.6454** | **Positive** | **0.2270** | **1.8694** |
| 21 | Positive | 3.8911 | 1.9976 | Positive | 0.6885 | 5.6713 |
| 22 | Negative | 0.2088 | 0.2645 | Negative | 0.0517 | 0.4255 |
| 23 | Negative | 0.2221 | 0.1648 | Negative | 0.0071 | 0.0585 |
| 24 | Negative | 0.1635 | 0.1754 | Negative | 0.0089 | 0.0729 |
| 25 | Negative | 0.2032 | 0.2628 | Negative | 0.0428 | 0.3526 |
| 26 | Positive | 3.9031 | 1.5984 | Positive | 1.3099 | 10.7900 |
| 27 | Negative | 0.2109 | 1.0731 | Negative | 0.1066 | 0.8777 |
| 28 | Negative | 0.2123 | 0.2391 | Negative | 0.0813 | 0.6697 |
| 29 | Negative | 0.1325 | 0.1466 | Negative | 0.0050 | 0.0412 |
| 30 | Positive | 3.7932 | 0.9603 | Positive | 0.6937 | 5.7142 |
| 31 | Positive | 2.3932 | 1.5027 | Positive | 0.2116 | 1.7430 |
| 32 | Negative | 0.2703 | 0.3878 | Negative | 0.1121 | 0.9234 |
| 33 | Negative | 0.2216 | 0.3375 | Negative | 0.0015 | 0.0124 |
| 34 | Negative | 3.6172 | 0.4099 | Negative | 0.0972 | 0.8007 |
| 35 | Positive | 3.1024 | 0.7545 | Positive | 0.6504 | 5.3575 |
| Cutoff | | | | | 0.1215 | |
| Negative control | | | | Negative | 0.0014 | 0.0111 |
| Positive control | | | | Positive | 2.0219 | 16.6544 |
| Positives = mean OD 450/cutoff >1 | | | | | | |

Appendix Table 3. SARS-CoV-2 Surrogate Virus Neutralization Test (sVNT) Kit results.

| Subject number | Mean OD sVNT assay | Mean / Postive CTRL | (1-Mean/Postive CTRL) | (%) Neutralizing capacity | Detectable SARS-CoV-2 neutralizing antibody | Mean RBD | Mean Spike | SPIKE / RBD seropostitive |
|---|---|---|---|---|---|---|---|---|
| 1 | 2.9965 | 1.3873 | -0.3873 | -38.73 | NO | 0.2157 | 0.2637 | Negative |
| 2 | 2.9837 | 1.3814 | -0.3814 | -38.14 | NO | 0.0813 | 0.1241 | Negative |
| 3 | 3.0597 | 1.4166 | -0.4166 | -41.66 | NO | 0.0847 | 0.3078 | Negative |
| 4 | 3.0404 | 1.4076 | -0.4076 | -40.76 | NO | 0.048 | 0.0532 | Negative |
| 5 | 2.8734 | 1.3303 | -0.3303 | -33.03 | NO | 0.08545 | 0.5203 | Negative |
| 6 | 2.9994 | 1.3886 | -0.3886 | -38.86 | NO | 0.06165 | 0.1983 | Negative |
| 7 | 3.0158 | 1.3962 | -0.3962 | -39.62 | NO | 0.0613 | 0.1358 | Negative |
| 8 | 2.8516 | 1.3202 | -0.3202 | -32.02 | NO | 0.03685 | 0.0361 | Negative |
| 9 | 2.2910 | 1.0607 | -0.0607 | -6.07 | NO | 3.74435 | 0.7881 | POSITIVE |
| 10 | 2.8065 | 1.2993 | -0.2993 | -29.93 | NO | 0.1912 | 0.2379 | Negative |
| 11 | 2.2671 | 1.0496 | -0.0496 | -4.96 | NO | 3.87495 | 0.4312 | Negative |
| 12 | 2.3722 | 1.0982 | -0.0982 | -9.82 | NO | 3.8917 | 0.428 | Negative |
| 13 | 2.5024 | 1.1585 | -0.1585 | -15.85 | NO | 3.8971 | 0.3964 | Negative |
| 14 | 2.5129 | 1.1634 | -0.1634 | -16.34 | NO | 3.08685 | 0.391 | Negative |
| 15 | 2.4956 | 1.1554 | -0.1554 | -15.54 | NO | 3.89305 | 0.9565 | POSITIVE |
| 16 | 2.5930 | 1.2005 | -0.2005 | -20.05 | NO | 3.85315 | 0.9306 | POSITIVE |
| 17 | 2.2649 | 1.0486 | -0.0486 | -4.86 | NO | 2.56455 | 0.7263 | POSITIVE |
| 18 | 2.5655 | 1.1878 | -0.1878 | -18.78 | NO | 3.8824 | 1.992 | POSITIVE |
| 19 | 2.2631 | 1.0477 | -0.0477 | -4.77 | NO | 3.47425 | 0.9039 | POSITIVE |
| 20 | 2.4560 | 1.1371 | -0.1371 | -13.71 | NO | 2.2409 | 0.6454 | Negative |
| 21 | 1.8539 | 0.8583 | 0.1417 | 14.17 | NO | 3.8911 | 1.9976 | POSITIVE |
| 22 | 2.8783 | 1.3326 | -0.3326 | -33.26 | NO | 0.2088 | 0.2645 | Negative |
| 23 | 2.8563 | 1.3224 | -0.3224 | -32.24 | NO | 0.2221 | 0.1648 | Negative |
| 24 | 2.6625 | 1.2326 | -0.2326 | -23.26 | NO | 0.1635 | 0.1754 | Negative |
| 25 | 2.7740 | 1.2843 | -0.2843 | -28.43 | NO | 0.20315 | 0.2628 | Negative |
| 26 | 2.4614 | 1.1396 | -0.1396 | -13.96 | NO | 3.90305 | 1.5984 | POSITIVE |
| 27 | 2.8388 | 1.3143 | -0.3143 | -31.43 | NO | 0.21085 | 1.0731 | Negative |
| 28 | 2.7149 | 1.2569 | -0.2569 | -25.69 | NO | 0.2123 | 0.2391 | Negative |
| 29 | 2.6724 | 1.2373 | -0.2373 | -23.73 | NO | 0.13245 | 0.1466 | Negative |
| 30 | 2.2980 | 1.0639 | -0.0639 | -6.39 | NO | 3.79315 | 0.9603 | POSITIVE |
| 31 | 1.9825 | 0.9178 | 0.0822 | 8.22 | NO | 2.39315 | 1.5027 | POSITIVE |
| 32 | 2.1600 | 1.0000 | 0.0000 | 0.00 | NO | 0.27025 | 0.3878 | Negative |
| 33 | 2.8539 | 1.3213 | -0.3213 | -32.13 | NO | 0.22155 | 0.3375 | Negative |
| 34 | 2.4904 | 1.1530 | -0.1530 | -15.30 | NO | 3.6172 | 0.4099 | Negative |

| | | | | | | | | |
|---|---|---|---|---|---|---|---|---|
| 35 | 2.3534 | 1.0895 | -0.0895 | -8.95 | NO | 3.10235 | 0.7545 | POSITIVE |
| Neg. CTRL | 2.5788 | 1.1939 | -0.1939 | | NO | | | |
| Pos. CTRL | 0.0863 | 0.0399 | 0.9601 | 96.01 | YES | | | |
| SARS-CoV-2 Neutralizing Antibody Standard serial dilution | | | | | | | | |
| 156.25 | 1.1358 | 0.5258 | 0.4742 | 47.42 | YES | | | |
| 312.5 | 0.5059 | 0.2342 | 0.7658 | 76.58 | YES | | | |
| 625 | 0.1636 | 0.0757 | 0.9243 | 92.43 | YES | | | |
| 1250 | 0.0687 | 0.0318 | 0.9682 | 96.82 | YES | | | |
| 2500 | 0.0490 | 0.0227 | 0.9773 | 97.73 | YES | | | |
| 5000 | 0.0465 | 0.0215 | 0.9785 | 97.85 | YES | | | |
| 10000 | 0.0517 | 0.0239 | 0.9761 | 97.61 | YES | | | |

Appendix Figure 1. Microneutralization of SARS-CoV-2 by pre-pandemic Cambodian sera

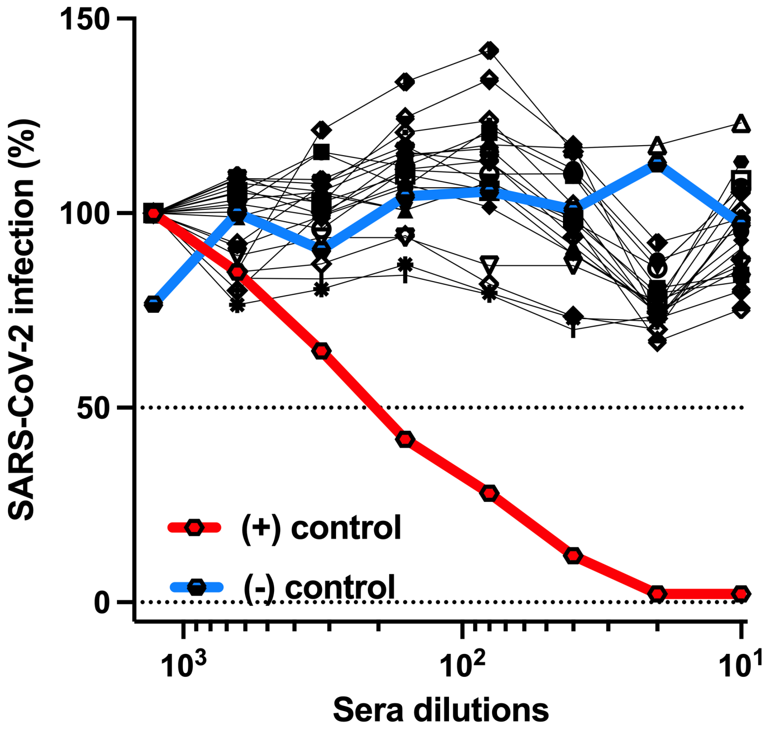

**Appendix Figure 1 Legend.** Twenty-one SARS-CoV-2 ELISA-positive Cambodian serum samples, negative controls (sera from US patients in 2014), and positive controls (sera from US patients who succumbed to SARS-CoV-2 infection in 2020) were heat-inactivated (56°C, 1hr), serially 2-fold diluted (1:10 to 1:1,280) in OptiPRO SFM (catalog no. 12309-019, Thermo Fisher) supplemented with 2mM L-Glutamine (catalog no. 25030-081, Thermo Fisher) and 1x Antibiotic-Antimycotic (catalog no. 15240062, Thermo Fisher). Diluted serum samples were mixed with an equal volume of SARS-CoV-2 diluted to 200 TCID50/25µl (USA-WA1/2020, catalog no. NR-52281, BEI resources) and incubated at room temperature for 1 hr. Fifty micro liter of the virus-plasma mixture was added in triplicate to Vero cells grown in a 96-well plate and incubated for 3 days at 37°C in a humidified incubator with 5% CO2. After incubation, media was removed and 200µl of 10% neutral buffered formalin (NBF) was added and incubated at room temperature for 30 min to inactivate the virus and fix the cells. After incubation, the plates were washed three times with wash buffer (0.05% Tween 20 in PBS), and 1:4,000 diluted SARS-CoV-2 nucleocapsid antibody (catalog no. 40143-R001, Sino Biological) was added to each well (50 µl per well). After incubation (room temperature, 1hr), the plates were washed 3 times, and 1:10,000 diluted horseradish peroxidase (HRP)-conjugated anti-rabbit IgG antibody (catalog no. 32460, Thermo Fisher) was added (100 µl per well), and the plates were incubated at room temperature for 1 hr. The plates were then washed 6 times followed by 30 min of room temperature incubation with HRP substrate solution (100 µl per well) prepared by adding a 10-mg o-phenylenediamine dihydrochloride tablet (catalog no. P8287, Millipore Sigma) to 20 ml of phosphate citrate buffer preparation (catalog no. P4922, Millipore Sigma). The reaction was stopped by adding 1 M sulfuric acid (100 µl per well), and the optical density was measured at 492 nm (OD492). To calculate percent infection, the optical density obtained with the lowest amount of serum (1:1,280) was used to set 100% infection for each serum sample.
